# A rapid review of equity considerations in large-scale testing campaigns during infectious disease epidemics

**DOI:** 10.1101/2021.02.22.21252205

**Authors:** Katarina Ost, Louise Duquesne, Claudia Duguay, Lola Traverson, Isadora Mathevet, Valéry Ridde, Kate Zinszer

**Affiliations:** University of Montreal, Montreal, Canada; University of Ottawa, Ottawa, Canada; Centre de recherche en santé publique, Montreal, Canada; CEPED, Institute for Research on Sustainable Development, IRD-Université de Paris, ERL INSERM SAGESUD, Paris, France

## Abstract

**Context:** Large-scale testing is an intervention that is instrumental for infectious disease control and a central tool for the COVID-19 pandemic. Our rapid review aimed to identify if and how equity has been considered in large-scale testing initiatives.

**Methods:** We searched Web of Science and PubMed in November 2020 and followed PRISMA recommendations for scoping reviews. Articles were analyzed using descriptive and thematic analysis.

**Results:** Our search resulted in 291 studies of which 41 were included for data extraction after full article screening. Most of the included articles (83%) reported on HIV-related screening programs, while the remaining programs focused on other sexually transmitted infections (n=3) or COVID-19 (n=4). None of the studies presented a formal definition of (in)equity in testing, however, 23 articles did indirectly include elements of equity in the program or intervention design, largely through the justification of their target population.

**Conclusion:** The studies included in our rapid review did not explicitly consider equity in their design or evaluation. It is imperative that equity is incorporated into the design of infectious disease testing programs and serves as an important reminder of how equity considerations are needed for SARS-CoV-2 testing and vaccination programs.

## 1. INTRODUCTION

It is essential to understand how COVID-19 testing campaigns are being offered in the current pandemic situation, in order to improve their equitable implementation. Racialized and marginalized communities have been disproportionately affected by COVID-19 (1–3) and improving equitable access to COVID-19 testing would be a vital step in reducing disease propagation (4). Large-scale testing is instrumental for surveillance, directly informing measures of prevention, control, and mitigation of infectious diseases (5–8). The goal of large-scale testing interventions is to reduce transmission rates through detection, treatment, isolation, and any other relevant control and prevention measures (9). Testing programs often act as a link to care and support programs, which should be provided equitably, based on risk of infection and disease burden (10). A proportionate universalism framework-based public health program would imply two components: a universal approach of support and services available to the population as a whole, accompanied by accessible targeted initiatives for those highly vulnerable and for those least likely to benefit from the universal program (11,12). When applied to COVID-19 testing initiatives, a proportionate universalism approach could include a universal program for SARS-CoV-2 testing with concerted efforts to reach vulnerable groups less able to access the universal testing programs.

Equity is defined by the “absence of systematic disparities in health or in major social determinants of health between groups with different levels of underlying social advantages/disadvantages’’ (13,14) whereas health inequities refer to “differences in health status or in the distribution of health resources between different population groups, arising from the social conditions in which people are born, grow, live, work and age. They are unfair, avoidable, and could be reduced by the right mix of government policies.” (15,16). This is an important concept for understanding the differences between (in)equity from the more general term (in)equality, two words that are often confused (17,18). Health inequalities refer to the uneven distribution of health or health resources (i.e. clinics, healthcare providers, disease tests, infrastructure, clinical material) in or between populations and it is primarily a descriptive term exempt from moral perspective (17–19). Social determinants of health play a key role in both inequality and inequity. Disparities in social determinants are found along social gradients (1) and are often avoidable as they result from deeply rooted social institutions, practices, and injustices (18). Thus, (in)equity is the politicized expression of (in)equality involving a moral commitment to social justice (18). In light of these fundamental differences, equity-sensitive public health interventions require measures of health and social determinants of health specific and sensitive to the health issue at hand (20). In our review we identify if and how equity has been considered in large-scale infectious disease testing initiatives. Identifying examples of (in)equity in these initiatives can help guide the design of large-scale testing campaigns for the COVID-19 pandemic.

## 2. METHODS

We chose to conduct a rapid review approach as it enabled us to synthesize, with rigor and in a relatively short period of time, the state of knowledge about our research objective (21,22). We have a detailed online protocol published elsewhere (23).

### 2.1 Research strategy

The research strategy was developed in consultation with librarians from the French National Research Institute for Sustainable Development (IRD) and the University of Montreal. We began our electronic database query in July 2020 on PubMed and Web of Science, and updated our search in November 2020 to better reflect the rapidly evolving state of COVID-19 literature. The following english and french key words were used to define our queries (Appendix 1) : “testing”, “mass testing”, “dépistage”, “screening”; “TB”, “tuberculosis”, “tuberculose”; “HIV”, “VIH”, “human immunodeficiency virus”; “COVID-19”, “SARS-CoV-2”, “coronavirus”; “design”, “planification”, “planning”; “equit*”, “equal*”, “inégalités”, “inégalités sociales en santé”, “ISS”, “social inequities in health”; “pandemi*”, “epidemic”, “outbreak”, “endemic”; “infectious disease”, “maladie infectieuse”.

We followed the PRISMA extension for scoping reviews (24). We used the Automated Text Classification of Empirical Records (ATCER) (25) tool to classify abstracts with an empirical degree ≥ 80. ATCER is a tool that automatically categorizes publications indexed in bibliographic databases into (a) empirical studies (>50), and (b) non-empirical work (<50) (25). We selected the ATCER threshold of ≥ 80 to reflect articles that were “highly empirical”, due to our objective of including studies with quantitative data such as program evaluation indicators. The inclusion criteria for articles were: i) a focus on an infectious disease, ii) description of the design portion of a testing or screening program, iii) published in English or in French, iv) had an empirical degree greater than 80 according to the ATCER tool, and v) published after 2010.

### 2.2 Selection of studies

All identified studies were imported from PubMed and Web of Science into Rayyan QCRI (26), a systematic review software, for screening of the titles, abstracts, and full texts. At least two of three involved reviewers (KO, LD, CD) independently assessed the relevance of titles and abstracts based on the inclusion and exclusion criteria. The second stage of review involved two of three reviewers independently identifying potentially relevant publications based on a full article review. Any discordance in the process was discussed among all reviewers and if no consensus was reached, an additional reviewer (LT) was consulted.

### 2.3 Study characteristics and data extraction

After independent full text screening was conducted by at least two reviewers, data from the retained articles were extracted and assessed. Extracted data included the following elements: characteristics (title, authors, year), context (country, disease addressed), and the consideration of health inequities or inequalities in the design of the intervention, the main results, and the discussion of the study. If the study considered health inequities, we extracted further information on which measures of equity were considered and if a specific tool or theoretical framework was used in the program design. Generally, a theoretical framework can be used to inform how a public health program is planned and what strategic and operational components were considered during the process of this planning (27). We considered equity (explicit or implicit) in the i) intervention rationale, ii) design, iii) choice of target population, and iv) final recommendations for future initiatives based on the PROGRESS-plus criteria (28). The PROGRESS Plus framework was developed and endorsed by the Campbell and Cochrane Equity Methods Group, in order to highlight a set of social determinants of health that drive variations in health outcomes and the inequalities among the social determinants of health gradient (28). The categories referred to by the acronym are: place of residence, race or ethnicity, occupation, gender, religion, educational level, socioeconomic status (SES), and social capital. Our goal, through the use of these tools, was to assess the presence or absence of the consideration of health inequities in the implementation or evaluation of testing programs.

We used the Mixed Methods Appraisal Tool (MMAT) (29), and specifically used the section on study methodology to apply a systematic definition to the included articles. The MMAT encompasses five study design categories: i) qualitative research, ii) randomized controlled trials, iii) non-randomized studies, iv) quantitative descriptive studies, and v) mixed methods studies.

In addition, we followed the Template for Intervention Description and Replication (TIDieR-PHP) checklist and guide to assess study coherence and program reporting and evaluation completeness (30). The 12-item checklist includes categories: (brief name, why, what (materials), what (procedure), who provided, how, where, when and how much, tailoring, modifications, how well (planned), how well (actual)) and is an extension of the CONSORT checklist.

## 3. RESULTS

### 3.1 Description of the studies

We initially identified 291 references with 41 peer reviewed studies being included in the review (Figure 1). According to MMAT classifications, the studies were a mixture of 2 randomized studies (31,32), 17 non-randomized studies (33–50), 18 descriptive quantitative studies (51–67), and 4 mixed-methods studies (68–71) (Table 1). Most of the studies (83%) reported on HIV-related screening programs, while the remaining studies focused on other sexually transmitted infections (n=3) and COVID-19 (n=4). The evaluated studies were implemented in North America (n=27), Europe (n=8), Africa (n=5), and Asia (n=1).

**TABLE 1:** Description of included articles (n=41)

| **Study** | **Country** | **Study Objectives** | **Study Design (MMAT) categorization** | **Findings** |
| --- | --- | --- | --- | --- |
| Nglazi et al. 2012, *An Incentivized HIV Counseling and Testing Program Targeting Hard-to-Reach Unemployed Men in Cape Town, South Africa* | South Africa | to investigate the impact of conditional cash transfer (CCTs) on HIV testing in men. | Quantitative non-randomized | incentivized mobile testing services may reach more previously untested men and significantly increase detection of HIV infection in men. |
| Bradshaw et al. 2018, *HIV/HCV/HBV testing in the emergency department: a feasibility and seroprevalence study* | United Kingdom | To identify: (i) how to embed sustainable testing for all three BBVs in the ED and (ii) the seroprevalence of HIV, HBV and HCV in this setting. | Quantitative descriptive | This program was found to be feasible and effective at case finding for BBVs, however there was a low testing rate and QI process had only moderate success, more strategies needed for sustained testing rates |
| Osorio et al. 2017, *Evaluation of opt-out inpatient HIV screening at an urban teaching hospital* | United States | To evaluate an opt-out inpatient HIV screening program administered with admission-orders written by physician house staff. These data were compared to the number of HIV tests and diagnoses as part of physician-ordered HIV testing (based on signs and symptoms) in the ED. | Quantitative non-randomized | First, opt-out inpatient HIV screening was associated with markedly lower per test positivity rates when compared to targeted testing in the ED, and these newly HIV diagnosed patients were not typically tested through physician-directed testing in the ED. Second, uptake of screening was limited when physicians were responsible for opt-out screening during routine care, with limited time resources seemingly the major barrier. |
| Porru et al. 2020, *Health Surveillance and Response to SARS-CoV-2 Mass Testing in Health* | Italy | To: (i) report HS data for SARS-CoV-2 with updated mass dx nasal and oropharyngeal swab tests in a very large cohort of symptomatic and asymptomatic HW, and (ii) learn | Quantitative non-randomized | Mass screening enabled isolation of HW, improved risk assessment allowed for close contacts of and infected HW to return to work in timely manner. |
| *Workers of a Large Italian Hospital in Verona, Veneto* |  | lessons regarding risk assessment, HS and other preventive strategies |  |  |
| Sicsic et al. 2016, *Impact of a primary care national policy on HIV screening in France: a longitudinal analysis between 2006 and 2013* | France | To (i) assess the impact of national policy on HIV screening launched in 2009 on a representative sample of the French general population aged 15-70 years and (ii) evaluate the independent effect of the frequency of visits to GPs. | Quantitative non-randomized | Mass screening policy coordinated by GPs had a significant impact on HIV testing in France. |
| Reiter et al. 2019, *Results of a Pilot Study of a Mail-Based Human Papillomavirus Self-Testing Program for Under screened Women From Appalachian Ohio* | United States | To implement a mail-based HPV self-testing program in Appalachian Ohio and examine how materials sent with the HPV self-test affect women's screening behaviors. | Quantitative Randomized | Mail-based HPV self-testing programs are a potentially promising strategy for reaching under-screened women in Appalachia. Efforts are needed to better understand how to optimize the success of such programs. |
| Musarandega et al. 2018, *Scaling up Pediatric HIV Testing by Incorporating Provider-Initiated HIV Testing Into all Child Health Services in Hurungwe District, Zimbabwe* | Zimbabwe | To describe how a program integrating HIV testing in pediatric health services was implemented and evaluates whether the program increased testing and identification of previously undiagnosed HIV-infected children below 5 years of age. | Quantitative descriptive | Integrating HIV testing into all pediatric health services is feasible and can assist in identifying HIV+ children who could be missed with current testing guidelines |
| Fuente et al. 2012, *Are Participants in a Street-Based HIV Testing Program Able to Perform Their Own Rapid Test and Interpret the Results?* | Spain | To evaluate the feasibility of HIV self-testing using a finger-stick whole-blood combo rapid test – including obtaining the sample and interpreting the results in conditions similar to those that would be found | Quantitative non-randomized | Blood-based self-testing with current technology is feasible in HIV-negative participants demanding the test and without prior training or supervision. Bearing in mind that it was conducted under difficult weather conditions and using a complex kit, over-the- |

|  |  | if this technology were available over the counter. |  | counter tests could be a feasible option to complement current diagnostic strategies. |
| --- | --- | --- | --- | --- |
| Ramirez-Avila et al. 2012, *Routine HIV Testing in Adolescents and Young Adults Presenting to an Outpatient Clinic in Durban, South Africa* | South Africa | To evaluate the number of adolescents (12–17 years) and young adults (18–24 years) who had HIV testing and the prevalence among those tested in Durban, South Africa. | Quantitative non-randomized | Although the HIV prevalence is high among youth participating in an adult outpatient clinic routine HIV program, the uptake of testing is low, especially among 12–17-year-old males. There is an urgent need to offer targeted, age-appropriate routine HIV testing to youth presenting to outpatient clinics in epidemic settings. |
| Barbee et al. 2016, *Effectiveness and Patient Acceptability of a Sexually Transmitted Infection Self-Testing Program in an HIV Care Setting* | United States | To evaluate the effectiveness and acceptability of a novel STI self-testing program in a large, urban HIV care clinic | Quantitative non-randomized | STI self-testing in HIV care settings significantly increases testing coverage and detection of gonorrhea and chlamydia, and the program is acceptable to patients. Additional interventions to increase syphilis screening rates are needed. |
| Go mez-Ayerbe et al. 2019, *Impact of a structured HIV testing program in a hospital emergency department and a primary care center* | Spain | To evaluate the impact of a structured HIV-testing intervention (DRIVE) in two clinical settings; hospital ED and primary care center in the same healthcare area, compared to HIV testing performed in routine clinical practice. | Quantitative non-randomized | Easy-to-implement structured intervention increased the absolute number of new HIV diagnoses and HIV tests compared to clinical practice. |
| Blackstock et al. 2010, *Evaluation of a Rapid HIV Testing* | United States | To describe the results of the dental clinic-based rapid HIV testing initiative. | Quantitative descriptive | Counselor-based rapid HIV testing program with linkage to specialized HIV care can be |
| *Initiative in an Urban, Hospital-Based Dental Clinic* |  |  |  | successfully integrated into the dental clinic setting. |
| Telford et al. 2020, *Preventing COVID-19 Outbreaks in Long-Term Care Facilities Through Preemptive Testing of Residents and Staff Members - Fulton County, Georgia, March-May 2020* | United States | To evaluate the timing at which mass testing took place in relation to the known presence of a COVID-19 infection and the resulting number of infections that occurred. | Quantitative non-randomized | Proactive testing of LTCF residents and staff members might prevent large COVID-19 outbreaks in LTCFs through early identification and timely infection prevention and control response. |
| Krueger et al. 2016, *Estimates of CDC-Funded and National HIV Diagnoses: A Comparison by Demographic and HIV-related Factors* | United States | To provide information about which groups disproportionately affected by HIV infection are being reached and which groups are underserved by CDC-funded HIV testing programs. | Quantitative non-randomized | CDC-funded HIV testing programs are reaching young people and African Americans. |
| Klein et al. 2014, *Missed Opportunities for Concurrent HIV-STD Testing in an Academic Emergency Department* | United States | To evaluate ED provider adherence to guidelines for concurrent HIV-sexually transmitted disease testing within an expanded HIV testing program and assess demographic and clinical factors associated with concurrent HIV-STD testing. | Quantitative descriptive | Concurrent HIV-STD testing in an academic ED remains low. Systematic interventions that remove the decision-making burden of ordering an HIV test from providers may increase HIV testing in this high-risk population of suspected STD patients. |
| Wachira, et al. 2013, *HIV Testing Uptake and Prevalence Among Adolescents and Adults in a Large Home-Based HIV Testing Program in Western Kenya* | Kenya | To (i) describe HIV testing uptake and prevalence in adolescents, younger adults, and older adults; and (ii) compare factors associated with HIV testing uptake and HIV prevalence among the 3 age groups. | Quantitative descriptive | Sociodemographic and socioeconomic factors were independently associated with HIV testing and prevalence among the age groups. Community-based treatment and prevention strategies will need to consider these factors. |
| Nunn et al. 2011, *Low Perceived Risk and High HIV Prevalence Among a Predominantly African American Population Participating in Philadelphia's Rapid HIV Testing Program* | United States | To assess actual and perceived risks in individuals undergoing rapid HIV testing in Philadelphia's program, analyze behavioral and demographic factors associated with newly diagnosed cases of HIV, and explain the important role of Philadelphia's rapid HIV testing program in diagnosing African Americans. | Quantitative descriptive | The Philadelphia experience suggests that routine, publicly financed HIV testing can play an important role in diagnosing HIV among high-risk individuals who do not otherwise have access to health services, particularly those who do not believe they are at risk for becoming infected. |
| Schumann et al. 2018, *Implementing a Standardized Social Networks Testing Strategy in a Low HIV Prevalence Jurisdiction* | United States | To describe Wisconsin's social networks testing program and outcomes. | Quantitative descriptive | Although social networks testing did not yield a higher new positivity rate compared to other testing strategies, it proved to be successful at reaching high risk individuals who may not otherwise engage in HIV testing. |
| Calderon et al. 2013, *Counselor-Based Rapid HIV Testing in Community Pharmacies* | United States | To examine the results of implementing counselor-based HIV testing and linkage to care components in five urban, NYC pharmacies located in communities highly affected by HIV, in areas with some of the highest rates of poverty in the United States. | Quantitative descriptive | Participants were satisfied with a counselor-based rapid HIV testing program in community-based pharmacies. Expansion of HIV screening initiatives into community pharmacies is one way to increase access to HIV testing for individuals who might not otherwise interact with the healthcare system |
| Lolekha et al. 2014, *Assessment of a couples HIV counseling and testing program for pregnant women and their partners in antenatal care (ANC) in 7 provinces, Thailand* | Thailand | To describe the pilot implementation of a CHTC program in ANC settings in 17 hospitals in 7 provinces in Thailand during 2009–2010; and to provide recommendations for improvement and scale-up of this program. | Mixed methods | Couples HIV testing and counseling implemented in antenatal care settings helps identify more HIV-positive men whose partners were negative than previous practice, with high acceptability among hospital staff. |
| Bischof et al. 2015, *Detecting HIV Among Persons Accompanying Patients to an Infectious Diseases Clinic* | United States | To implement a program providing free rapid HIV testing as a point-of-care (POC) screening tool in the University of North Carolina (UNC) ID clinic, targeting individuals accompanying HIV-infected patients to their appointments. Notably, the program was developed and implemented without any dedicated funding, but used existing infrastructure and staff. | Mixed methods | The high positivity and confirmatory HIV rates in our program confirm that the provision of rapid HIV testing in an ID clinic capitalizes on missed opportunities among an at-risk population and allows immediate linkage to care |
| Hsieh, et al. 2017, *Factors associated with patients who prefer HIV self-testing over health professional testing in an emergency department-based rapid HIV screening program* | United Kingdom | To determine which factors were associated with patients who were more likely to accept kiosk-facilitated self-testing by exploring socio-demographic information, computer/kiosk technology experience, behavior patterns, and ED visit characteristics compared to those who chose conventional HIV testing by trained clinical staff. | Quantitative descriptive | Approximately 50% of ED patients who accepted HIV testing chose to perform self-testing by following the instructions provided by a kiosk. Preference of self-testing was positively correlated with young age but negatively correlated with an ED primary diagnosis of infectious disease. With 50% of patients choosing conventional healthcare professional testing over self-testing in the ED, our findings indicate that ED patient self-testing for HIV is likely a complementary testing approach to the current, traditional format. |
| Qvist et al. 2014, *High Linkage to Care in a Community-Based Rapid HIV Testing and Counseling Project Among Men Who Have Sex With Men in Copenhagen* | United States | To evaluate a community-based HIV testing program for its capacity to reach men who have sex with men (MSM) and successfully refer HIV-positive patients to treatment. | Quantitative descriptive | Easily accessible, community walk-in clinics and targeted testing in high-risk settings are convenient for populations of MSM less likely to seek out the established health care system. Check-point diagnosed 37 new HIV cases, posed no barrier to successful link-age to care, was noninferior in quickly reducing community viral load, was cost-effective, reached younger MSM, and proved an ideal platform for trying out new interventions and test forms, which conventional health care providers have not yet embraced. |
| Cowan et al. 2018, *Bundled HIV and Hepatitis C Testing in the Emergency Department: A Randomized Controlled Trial* | United States | To integrate rapid HCV testing into an established HIV testing and counseling program to evaluate the effect of rapid, bundled screening on HIV-test acceptance rate. | Quantitative Randomized | Integrating bundled, rapid HCV/HIV testing into an established HIV testing program did not significantly impact HIV test acceptance. Future screening efforts for HCV could be integrated into current HIV testing models to target high-risk cohorts. |
| Williams-Roberts et al. 2010, *Frequent HIV testing among participants of a routine HIV testing program* | United States | To determine the characteristics of those more likely to undergo frequent (HIV) testing. | Quantitative descriptive | We found that providers need to strengthen practices to identify persons who have had multiple HIV tests and provide enhanced behavioral interventions for those with persistent risks. This might mean referral to other prevention and support services in order to effect sustainable risk reduction. The results also suggest that some risk behaviors (i.e. injection drug use, MSM and multiple sexual partners) are appropriately recognized as markers for more frequent HIV testing |
| Harmon et al. 2014, *Integrating Routine HIV Screening into a Primary Care Setting in Rural North Carolina* | United States | To (i) increase HIV testing in the Henderson community and surrounding rural area by integrating rapid HIV testing into the primary care setting; and (ii) to examine the relationship between sociodemographic variables and acceptance of HIV testing | Quantitative non-randomized | Early implementation of the NACHC model in our setting posed challenges in terms of time involved in initial planning, consistent data collection and reporting, and patient flow. In spite of these challenges, 100 patients were screened for HIV infection who might not have been screened otherwise, and they were given HIV risk reduction handouts after testing, an education intervention that may raise awareness and lead to behavior changes. Younger patients were more likely to undergo testing. The majority of patients who were tested, African American women, represented a high-risk group in North Carolina and the South, and yet African American MSM, those with the highest risk, were underrepresented in our sample. |
| Scognamiglio et al. 2018, *HIV rapid testing in community and outreach sites: results of a nationwide demonstration project in Italy* | Italy | To conduct a national demonstration project on the HIV rapid test offered outside the formal health facilities in collaboration with six NGOs in order to assess feasibility and potential yields of CBVCT implementation in different settings. This article reports the main finding of this project. | Quantitative non-randomized | Our project showed that the offering of an HIV rapid testing program in community and outreach settings in Italy is feasible and that it may reach people who have never been tested before, while having a significant yield in terms of new HIV diagnoses as well. |
| Spaulding et al. 2016, *Establishing an HIV Screening Program Led by Staff Nurses in a County Jail* | United states | To describe how a routine, opt-out, rapid HIV testing program has resulted in a substantial number of new HIV diagnoses and linkages to care. | Quantitative descriptive | A nurse-led, rapid HIV testing model successfully identified new HIV diagnoses. The testing program substantially decreased the number of persons who are HIV-infected but unaware of their status and promoted linkage to care. |
| Beckwith et al. 2011, *An Evaluation of a Routine opt-out Rapid HIV Testing Program in a Rhode Island Jail* | United states | To (i) assess the feasibility of rapid HIV testing within the jail as an alternative testing strategy; (ii) examine the rapid HIV testing pilot program; and (iii) explore provider and institutional stakeholder perspectives about the rapid HIV testing program. | Mixed methods | There was overwhelming support for the oral fluid rapid HIV test. Correctional staff reported improved inmate processing due to the elimination of phlebotomy required with conventional HIV testing. Delivering negative rapid HIV test results in real-time during the jail intake process remained a challenge but completion of confirmatory testing among those with reactive rapid tests was possible. Rapid HIV testing using oral specimens in the RIDOC jail was feasible and preferred by correctional staff. |
| Myers et al. 2015, *Uptake and Outcomes Associated with Implementation of a Community-Based Voluntary HIV Counseling and* | Antigua and Barbuda | To (i) describe the design and implementation of the CBVCT program in A&B; (ii) assess and describe associations between testers' demographic and risk characteristics and the receipt of a positive HIV test result; and (iii) | Quantitative non-randomized | The CBVCT model can be successfully implemented in Eastern Caribbean. Demographic differences persist in HIV testing, risk behavior, and infection among vulnerable populations and should be considered in HIV prevention intervention design. |
| *Testing Program in Antigua and Barbuda* |  | assess and describe associations between these same characteristics and consistent condom use, a proven-effective prevention strategy. |  |  |
| Sitlinger et al. 2011, *Preliminary Program Evaluation of Emergency Department HIV Prevention Counseling* | United States of America | To conduct a complementary series of investigations, using survey and chart review methodology to explore (i) patient perception and feasibility of prevention counseling in the ED, (ii) patient perceptions of the need to link prevention counseling with testing, and (iii) potential effectiveness of prevention counseling provided in conjunction with ED-based HIV testing. | Mixed methods | Participants in the ED prevention counseling and testing program considered counseling acceptable and useful, though not required. Given adequate resources, prevention counseling can be provided in the ED, but it is unlikely that all patients benefit. |
| Matulionyt et al. 2019, *Routine HIV testing program in the University Infectious Diseases Centre in Lithuania: a four-year analysis* | Lithuania | To (i) evaluate the feasibility and clinical effectiveness of fully integrated non targeted opt-out routine rapid HIV testing program; and (ii) determine whether routine testing in an urban teaching infectious disease hospital was associated with identification of more patients with newly diagnosed HIV infection than targeted rapid HIV testing. | Quantitative descriptive | Routine HIV testing in admissions of infectious diseases is acceptable, feasible, sustainable and clinically effective. Compared to targeted testing, routine testing helped to discover more patients in earlier stages and those with heterosexually transmitted HIV infection. |
| Lederman et al. 2020, *Opt-out Testing Pilot for Sexually Transmitted Infections Among Immigrant Detainees at 2 Immigration and Customs Enforcement Health Service Corps-* | United States | To explore the feasibility of implementing an opt-out STI screening program to inform future Performance-Based National Detention Standards guidance and practices in a subset of detention facilities in which we have direct visibility over patient care (ICE | Quantitative non-randomized | STIs are prevalent among immigrant detainees. A routine screening program is feasible if operational aspects are carefully considered and would provide counseling, education, and treatment for this vulnerable population. |
| *Staffed Detention Facilities, 2018* |  | Health Service Corps [IHSC]–staffed facilities). |  |  |
| Ndege et al. 2015, *HIV Prevalence and Antenatal Care Attendance among Pregnant Women in a Large Home-Based HIV Counseling and Testing Program in Western Kenya* | Kenya | To describe population-based uptake of and factors associated with HIV testing and HIV prevalence among pregnant women in a large-scale home-based HIV counseling and testing (HBCT) program in western Kenya. | Quantitative non-randomized | Pregnant women who had never attended ANC were about 6 times more likely to newly test HIV-positive compared to those who had attended ANC, suggesting that the cascade of services for prevention of mother-to-child HIV transmission should optimally begin at the home and village level if elimination of perinatal HIV transmission is to be achieved. |
| Imbert et al. 2020, *Coronavirus Disease 2019 (COVID-19) Outbreak in a San Francisco Homeless Shelter* | United States | To describe the lessons learned from our public health response to a COVID-19 outbreak that occurred. | Quantitative descriptive | This outbreak demonstrates the limited utility of case investigation, person-based contact tracing, and symptom screening, and the benefits of mass testing in outbreak response. Cases were widely distributed throughout the shelter, reinforcing the risks of congregate living and highly populated shelters without capacity for social distancing. Identification of cases through expanded testing should be done in conjunction with a clear strategy for isolating and quarantining confirmed positives and their close contacts. |
| Hubbard et al. 2020, *#Testathome: Implementing 2 Phases of a HIV Self-Testing Program Through Community-Based Organization Partnerships in New York City* | United States | To (i) pilot distribution of free HIVSTs among priority populations disproportionately impacted by HIV in NYC, (ii) describe the program model, including feasibility of partnering with CBOs, as well as program reach and key outcomes. | Quantitative descriptive | Through 2 phases of implementation, this innovative partnership was able to reach and offer HIVSTs to priority populations across NYC. This program supports the feasibility of distributing HIVSTs through close partnerships with diverse organizations. |
| White et al. 2010, *A Comparison of HIV Testing in the* | United States | To compare the results of HIV screening and diagnostic HIV testing in patients triaged to the ED | Quantitative non-randomized | Although the yield of HIV testing is greater among ED patients, urgent care (UC) patients |
| *Emergency Department and Urgent Care* |  | compared with those triaged to an affiliated UC. |  | are diagnosed at a less advanced stage of illness. |
| Baggett et al. 2020, *Addressing COVID-19 Among People Experiencing Homelessness: Description, Adaptation, and Early Findings of a Multiagency Response in Boston* | United States | To create a partnership between Boston Health Care for the Homeless Program (BHCHP), and city & state public health agencies, municipal leaders, and homeless service providers to proactively develop a comprehensive COVID-19 response model for homeless and marginally housed people in Boston. | Quantitative descriptive | During the first 6 weeks of operation, 429 of 1297 (33.1%) tested people were positive for COVID-19; of these, 395 people were experiencing homelessness at the time of testing, representing about 10% of the homeless adult population in Boston. Universal testing, as resources permit, is a focal point of ongoing efforts to mitigate the effect of COVID-19 on this vulnerable group of people. |
| Haines et al. 2011, *Pediatric Emergency Department-Based Rapid HIV Testing Adolescent Attitudes and Preferences* | United States | To describe adolescent attitudes and preferences toward rapid HIV testing in a Pediatric Emergency Department. | Quantitative descriptive | This study offers valuable new insights into adolescent attitudes and preferences for rapid HIV testing in a PED. Adolescents gave high ratings for the location, testing, and counseling process. Our data support the importance of structured counseling, which is contrary to current published perspectives of counseling efficacy. In addition, we found that the PED was a highly preferred location for rapid HIV testing, which supports the need for increased development of prevention and testing programs in this setting. |
| Miller et al. 2017, *Evaluating Testing Strategies for Identifying Youths With HIV Infection and Linking Youths to Biomedical and Other Prevention Services* | United States | To (i) evaluate a multisite HIV testing program designed to encourage localized HIV testing programs focused on self-identified sexual minority males (especially those of color) aged 13 to 24 years; and (ii) link youths to appropriate prevention services after receipt of their test results. | Quantitative non-randomized | The findings suggest that community-based targeted approaches to HIV testing are more effective than universal screening for reaching young sexual minority males (especially males of color), identifying previously undiagnosed HIV-positive youths, and linking HIV-negative youths to relevant prevention services. Targeted, community-based HIV testing strategies hold promise as a scalable and effective means to identify high-risk youths who are unaware of their HIV status. |
| Whalen et al. 2018,<br>*Translation of Public Health Theory into Nursing Practice: Optimization of a Nurse-Driven HIV Testing Program in the Emergency Department* | United States | To evaluate the impact of a focused emergency nurse partnership with a long-standing HIV testing program, by analyzing a successive series of nurse-driven strategies focused on optimizing rates of HIV testing. | Quantitative non-randomized | The emergency department is a prime location for public health interventions such as HIV screening. To incorporate these initiatives, strong nursing engagement and leveraging existing resources is key to success. In addition, true sustainability requires integration into the clinical workflow, including optimizing the 24-hour nature of the emergency department to ensure screening reaches all parts of the population. |

The main type of study setting among the included articles were hospitals (n=16) and clinic-based (n=10). Nine of the 16 hospital-based studies were in emergency departments (31,35,49,50,53,55,56,59,71), 6 were hospital inpatient or outpatient-based (43,45,60,61,69,70), and 1 was directed at hospital healthcare workers (44). Of the 10 clinic-based studies, 7 occurred in HIV/STI testing centers, public health clinics, or community health centers (36,39,42,62– 64,72), 2 were in general practitioner offices (35,47), and 1 was in an urgent care clinic (67). For participant recruitment, 13 studies recruited participants outside of healthcare testing settings, such as: prison-settings (38,65,68), community-based or mobile clinic (34,40,42,46), dental or pharmacy settings (52,54), homeless shelters (51,58), residential homes (48), and multiple sites and settings (37). Most studies (n=38) focused on a combination of adults over the age of 18 and sexually active individuals over the age of 13 (31,32,34–40,42–50,52–54,56–60,62–72). Two articles investigated pediatric populations (55,61), with one other study focused on newborns and their mothers (41) and one on the elderly (48).

### 3.2 Consideration of Health Equity, PROGRESS-Plus: Measures of (In)Equity

Measures of equity/inequity were assessed based on the PROGRESS-Plus framework. None of the 41 included studies evaluated whether the intervention reduced health inequity or inequalities as a study objective, nor did they include a formal definition of equity/inequity (or framework). Elements of health equity were indirectly addressed in 23 studies (Table 2), largely through the use of targeted interventions. Of these 23 studies, 20 included at least one category of PROGRESS-plus in the justification, methods, results or discussion of the paper, while only 8 of the 23 articles alluding to health equity addressed a PROGRESS-plus category in all relevant sections of their article (Table 2): from the introduction to the discussion and conclusion. The target populations were most often described as disadvantaged subgroups of the population who bore the largest burden of disease and/or have the least access to health care. For example, four articles, two of which focused on HIV and/or STI testing and two on COVID-19, all targeted their programs towards four different “high risk” populations. For the HIV and/or STI testing programs, one program (72)targeted men based on their sexual orientation whereas another program (62) focused on race and ethnicity. For the COVID-19 testing programs, one intervention (51) focused on those experiencing homelessness, whereas another was based in a long-term care facility (48).

**TABLE 2:**
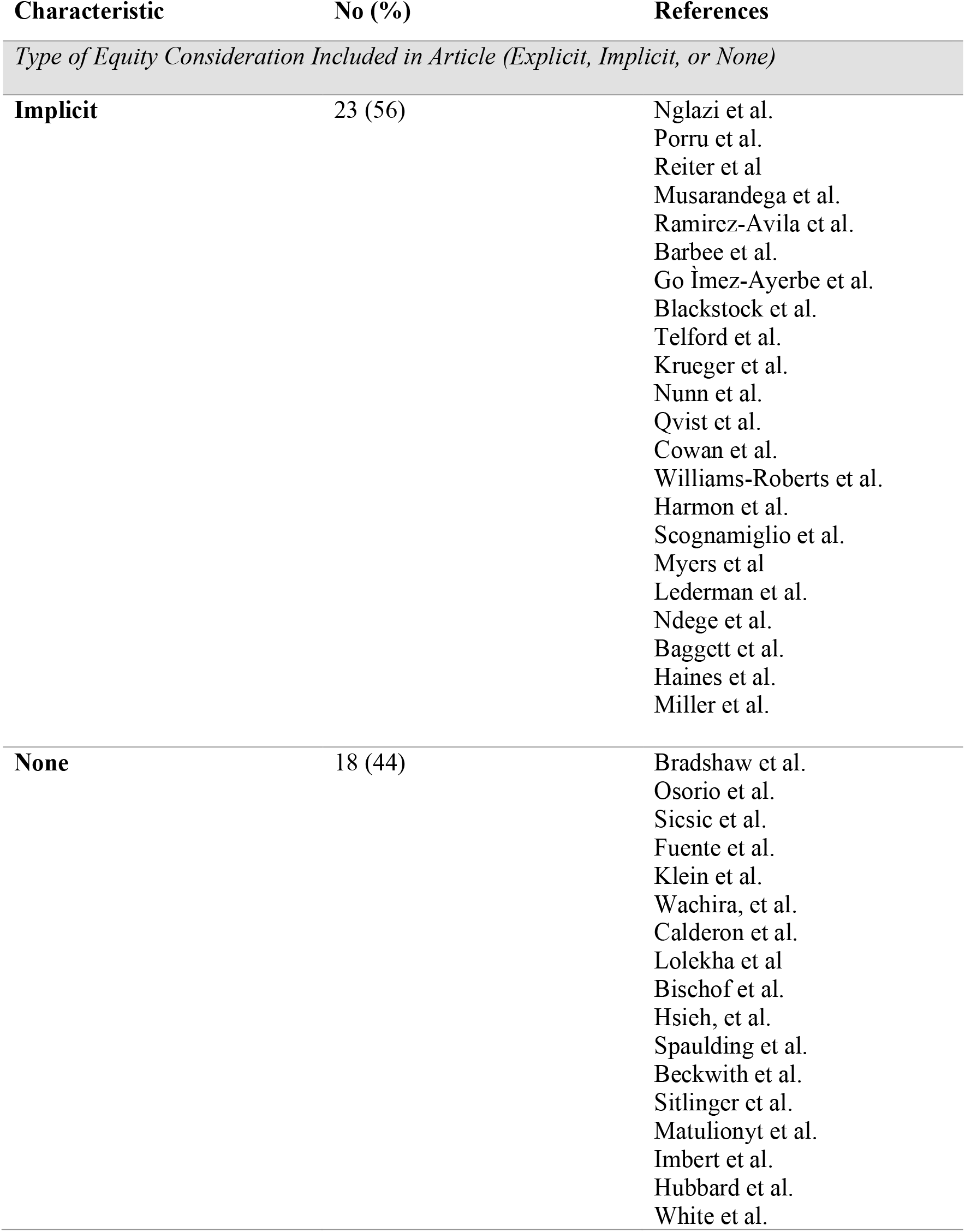
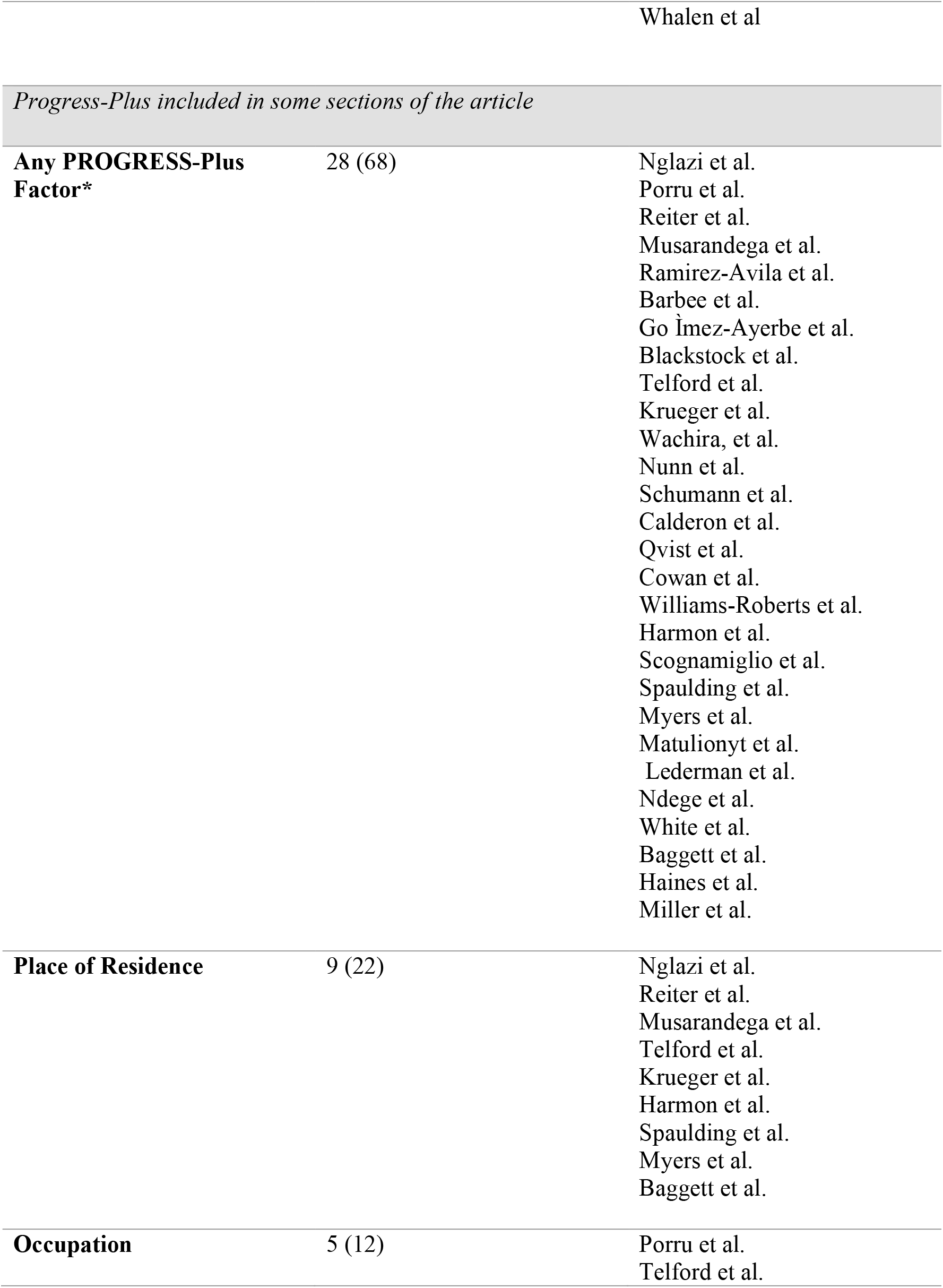
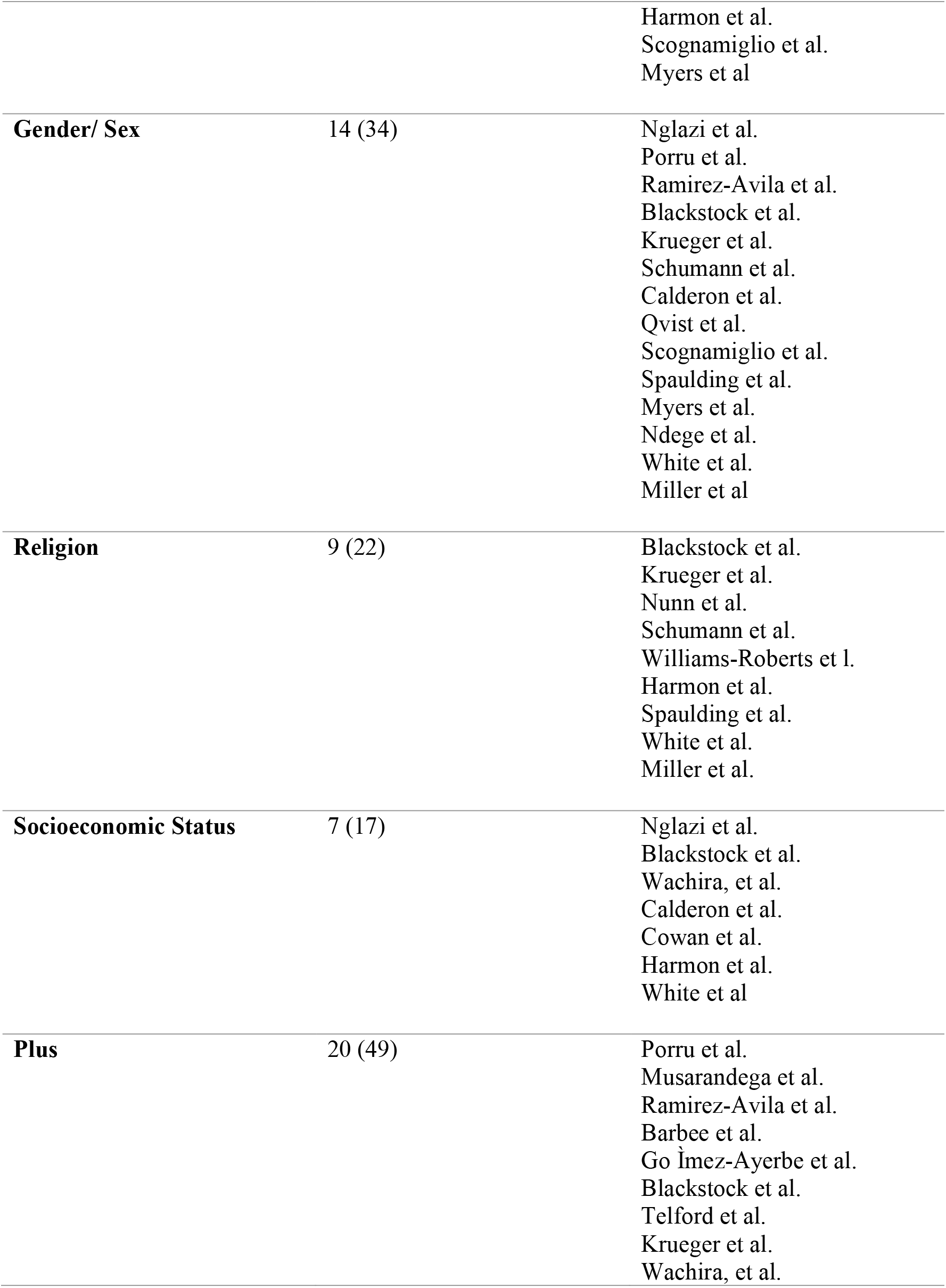
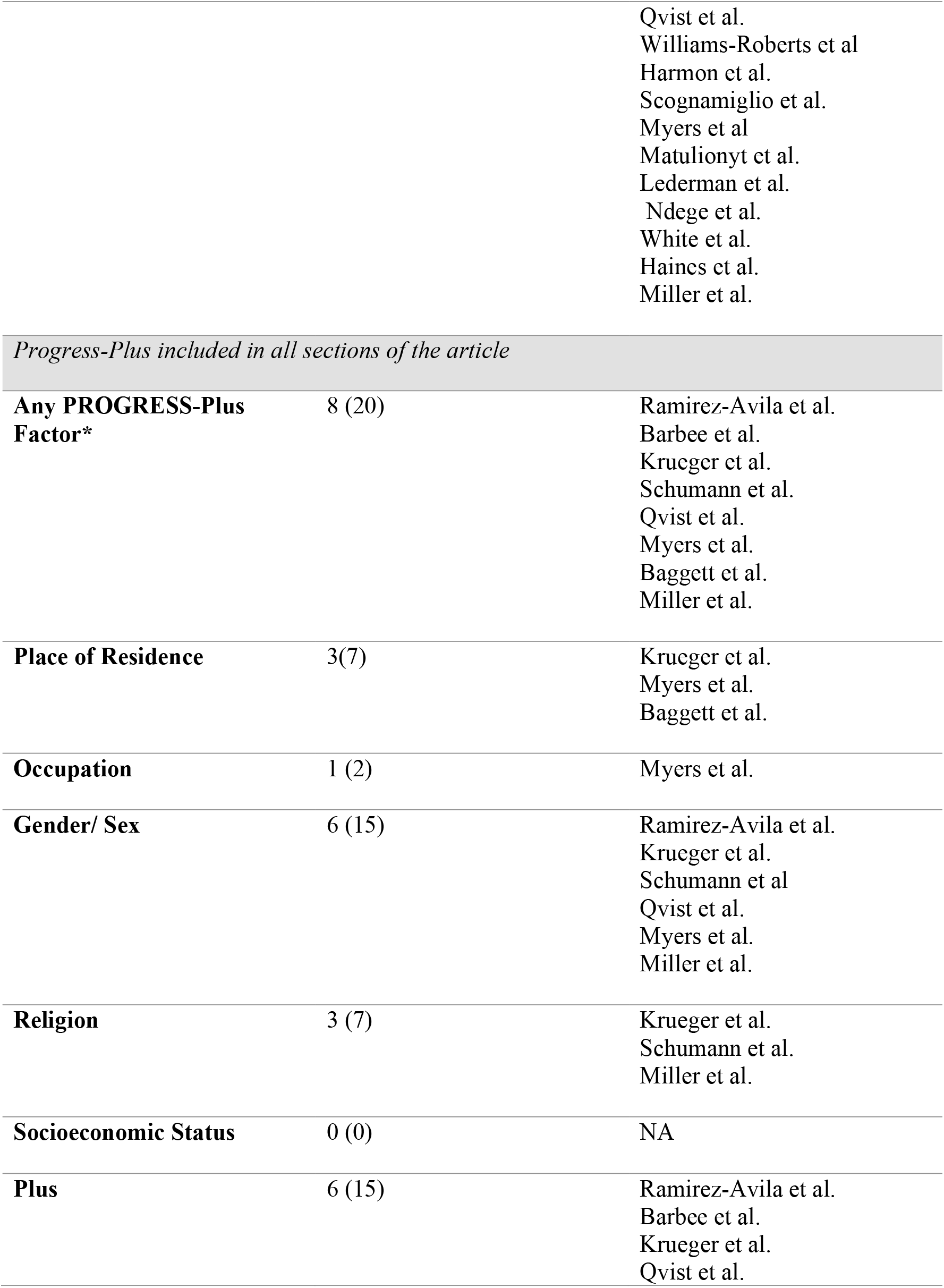
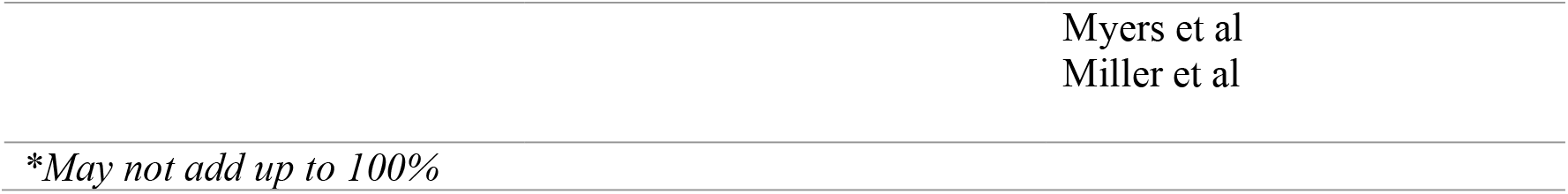
Results of equity appraisal using concepts of implicit/explicitly defined equity and PROGRESS-plus categories

## 4. DISCUSSION

This rapid review largely featured articles addressing HIV testing programs, and/or testing programs of infectious disease in North America in formal healthcare settings such as clinics or hospitals. None of the 41 studies included in this rapid review examined health equity in their interventions, however, 8 studies did consistently include elements of equity in their testing intervention, without the inclusion of any formal measurement of, or framework specifically implemented in order to address health equity. Specifically, most (n=6) of these 8 articles considered the PROGRESS-plus categories of gender and/or sex.

To our knowledge, no prior studies have examined the equitable access, delivery, or design of COVID-19 testing programs. This review shows that implicit measures of equity have been implemented through targeting COVID-19 testing programs to high-risk populations such as health care workers (44), people living in homeless shelters (51,58,73) and long-term care facilities (48). It is largely recognized that targeted actions towards specific communities or groups imply labelling, thus stigma, and increases the risk of missing numerous infections in particularly vulnerable population groups (11,12,74,75). The current pandemic is exacerbating health inequities and testing programs need to be designed accordingly to address these inequities, which are also central to mitigating disease spread (76–79). An example of an approach that could be used in combination with existing testing infrastructure to increase equitable access to COVID-19 testing consists of the deployment of rapid antigen testing kits for in-home testing for those with faced with mobility or geographic restrictions or work and/or family obligations that do not easily coincide with testing program schedules (3).

Numerous tools exist to support incorporating equity values within public health interventions and programs (80–82). These tools should be used both during the design, implementation, and evaluation process, as the link between equity intention and action remains challenging (83). TIDieR-PHP was developed to enable consistent reporting of Population Health and Policy (PHP) interventions to promote transparency and transferability of findings to diverse settings. PHP interventions are crucial to addressing disparities in social and economic determinants of health (30). This tool, when used in combination with the PROGRESS-plus framework, should ensure that researchers and practitioners are thinking critically about what health inequities they are addressing, how they are being measured and considered consistently throughout the program design, implementation, and evaluation process. To ensure that public health interventions are developed and implemented around the concept of equity, strong health policies and educational goals of healthcare professionals need to be on the public political and economic agenda (84).

**Recommendations towards improving the inclusion of health equity in large-scale testing interventions**

- Use tools such as PROGRESS-Plus framework to ensure explicit inclusion of health equity when in the process of designing, implementing, and/or evaluating interventions.
- Promote the use of TIDIER-PHP to systematically review public health programs and promote replicability of existing equitable programs to other settings.

### 4.1 Limitations

Given our objective to include studies with empirical results and also to conduct the review in a timely manner, we chose an ATCER threshold of 80. This may have limited our findings and the generalizability of the results. We recommend that a full scoping review be conducted on this topic to further investigate important trends on the incorporation of health equity into infectious disease testing programs.

### 4.2 Conclusion

The results of this rapid review highlight the overall lack of consideration of equity in the design of large-scale testing interventions. This is a particularly concerning issue as social and economic inequities continue to be exacerbated by COVID-19 and there has not been any research to date that discusses how COVID-19 testing programs have been designed with equity in mind (1–3). To achieve equity in testing and to optimize the role of testing in disease prevention and control, strategies should ensure that the probability of being tested is proportionate to the risk of being affected by the disease (85,86). We urge practitioners, decision makers, and researchers to explicitly include equity measures when designing and implementing COVID-19 large-scale testing interventions, which should also be considered in COVID-19 vaccination programs.

## Data Availability

All rapid review search criteria, databases, and keywords are listed in the manuscript or appendix.

## 5. ACKNOWLEDGEMENTS

The authors would like to thank Laurence Goury, librarian at the IRD, and Julie Desnoyers, librarian at the University of Montreal, for their advice regarding the search strategy and the queries on the bibliographic databases.

## 6. FUNDING SOURCES, CONFLICTS OF INTEREST

This work was supported by The Fonds de recherche du Québec – Santé (FRQS) [ZINKA170, 2020]; CIHR Canadian 2019 Novel Coronavirus Rapid Research Competition [440254]; and the French National Research Agency (ANR) (ANR-20-COVI-000). Authors have no conflicts of interest to declare.

## APPENDIX 1 PRISMA diagram for selection of articles for data extraction

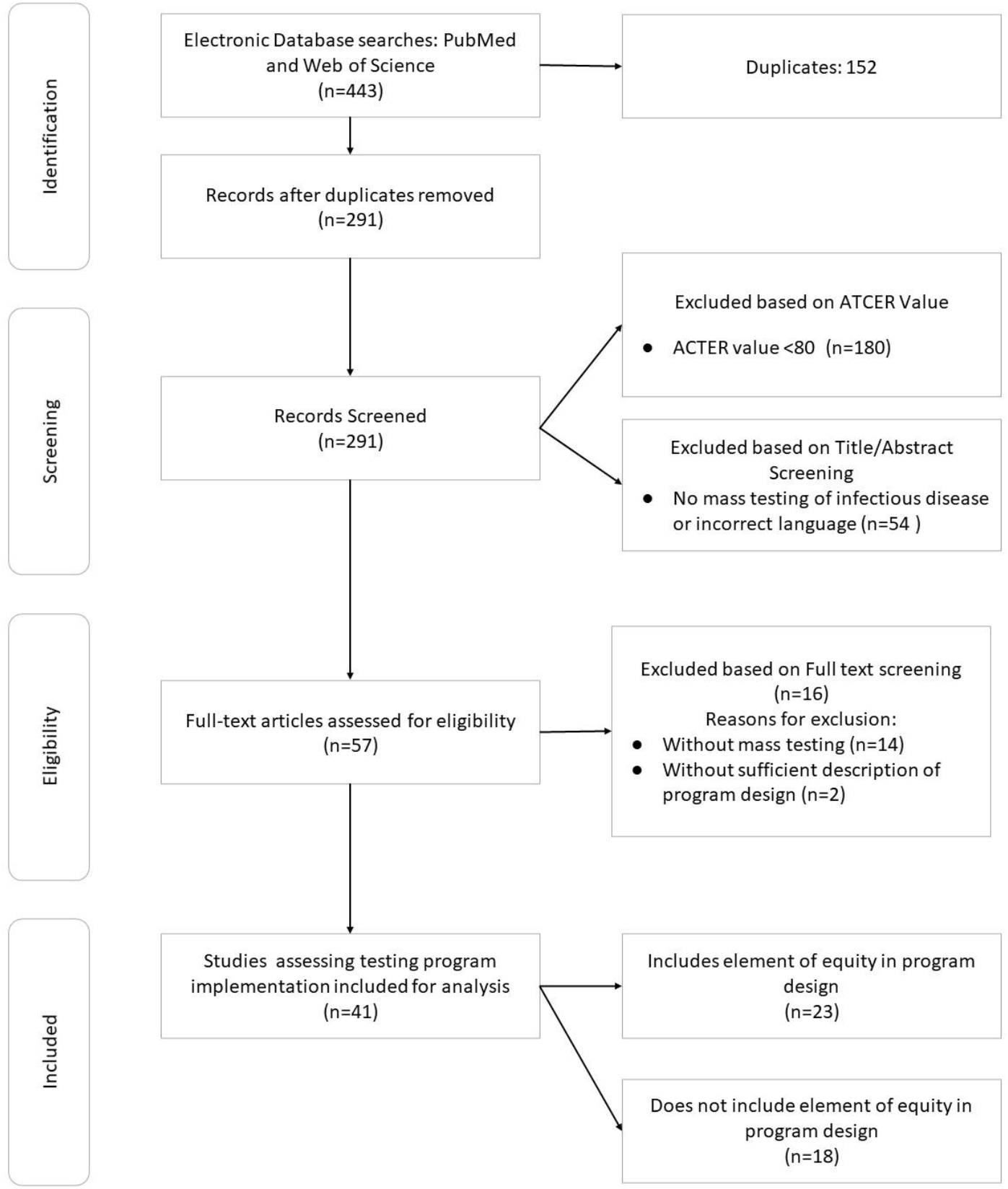

## APPENDIX 2 Queries in the electronic databases PubMed and Web of Science (filled in on 15/11/2020)

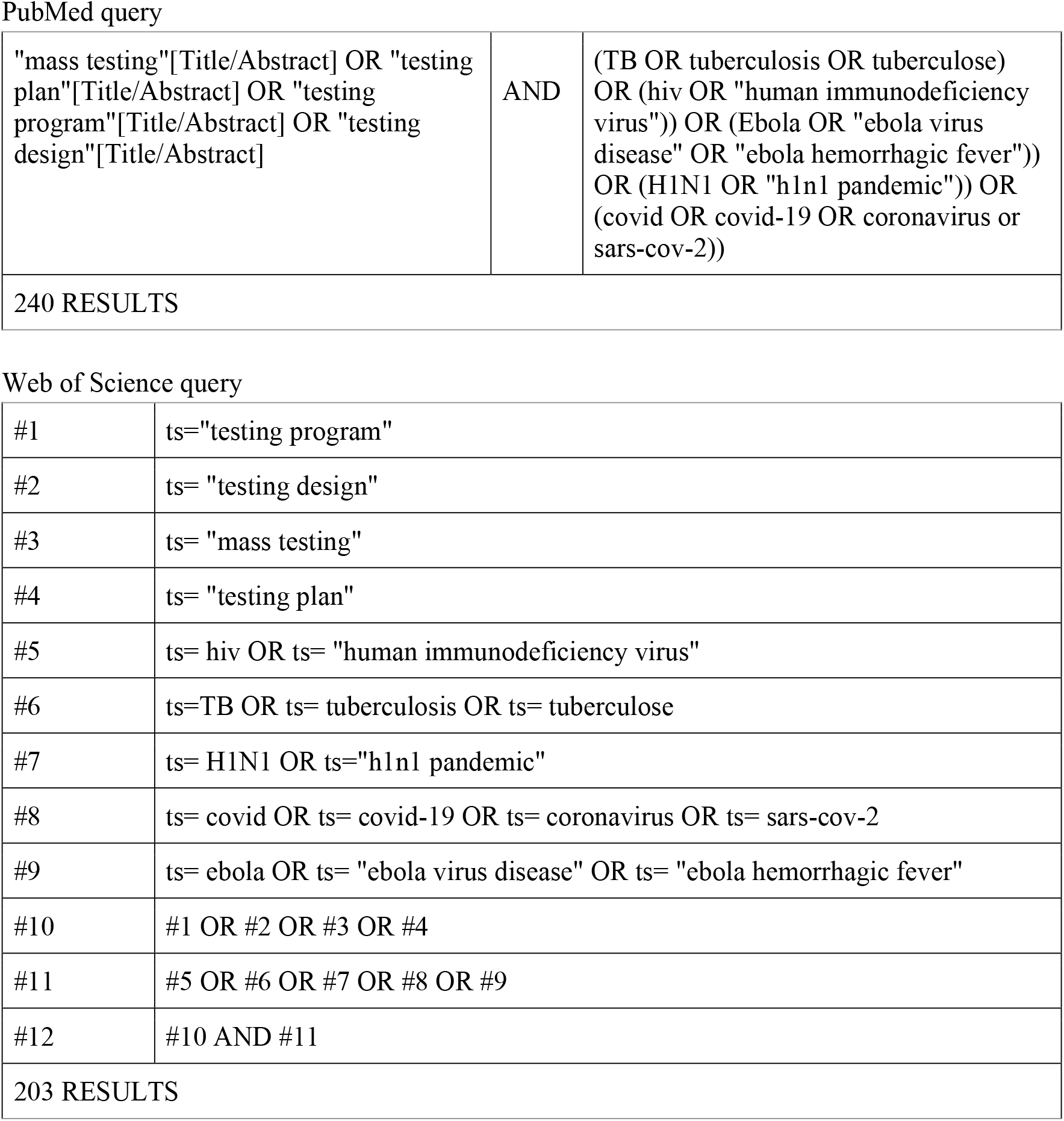

|  |  |  |
| --- | --- | --- |
| "mass testing"[Title/Abstract] OR "testing plan"[Title/Abstract] OR "testing program"[Title/Abstract] OR "testing design"[Title/Abstract] | AND | (TB OR tuberculosis OR tuberculose) OR (hiv OR "human immunodeficiency virus")) OR (Ebola OR "ebola virus disease" OR "ebola hemorrhagic fever")) OR (H1N1 OR "h1n1 pandemic")) OR (covid OR covid-19 OR coronavirus or sars-cov-2)) |
| 240 RESULTS |  |  |

Web of Science query
|  |  |
| --- | --- |
| #1 | ts="testing program" |
| #2 | ts= "testing design" |
| #3 | ts= "mass testing" |
| #4 | ts= "testing plan" |
| #5 | ts= hiv OR ts= "human immunodeficiency virus" |
| #6 | ts=TB OR ts= tuberculosis OR ts= tuberculose |
| #7 | ts= H1N1 OR ts="h1n1 pandemic" |
| #8 | ts= covid OR ts= covid-19 OR ts= coronavirus OR ts= sars-cov-2 |
| #9 | ts= ebola OR ts= "ebola virus disease" OR ts= "ebola hemorrhagic fever" |
| #10 | #1 OR #2 OR #3 OR #4 |
| #11 | #5 OR #6 OR #7 OR #8 OR #9 |
| #12 | #10 AND #11 |
| 203 RESULTS |  |

## Notes

### Competing Interest Statement

The authors have declared no competing interest.

### Funding Statement

This work was supported by The Fonds de recherche du Quebec: Sante (FRQS) [ZINKA170, 2020]; CIHR Canadian 2019 Novel Coronavirus Rapid Research Competition [440254]; and the French National Research Agency (ANR) (ANR-20-COVI-000). Authors have no conflicts of interest to declare.

### Author Declarations

Rapid review of peer-reviewed literature, no IRB approval required.

